# LONG TERM NEUROPSYCHOLOGICAL OUTCOME AND EFFECT OF COGNITIVE RESERVE IN RETIRED ATHLETES

**DOI:** 10.1101/2024.11.05.24316739

**Authors:** B-K. Dewar, J. Murray, H. Pedder, M. Turner

## Abstract

**Objectives:** Cognitive reserve is hypothesised as a protective process against cognitive impairment arising from brain injury. Different life experiences provide a shield against the effects of brain changes to modulate or delay the expression of cognitive impairment. The current study investigated the long-term neuropsychological outcome of retired, predominantly equestrian, athletes.

**Methods:** Neuropsychological performance of participants with a history of concussion was compared to age-matched controls using Principle Components Analysis (PCA) regression, adjusted for participant characteristics. Premorbid function was used as an index of cognitive reserve.

**Results:** Three components were identified that explained greater than 95 percent of the variance in 14 neuropsychology tests; the first component (PC1) explained 87 percent of the variance. A regression model on PC1 identified an association between concussion and PC1, with lower scores in concussed participants with lower premorbid function.

**Conclusions:** A history of concussion is associated with lower neuropsychological performance, and this is more pronounced in individuals with lower premorbid functioning, an indicator of cognitive reserve.

*What is already known on this topic:* There is a need to clarify long term neuropsychological outcome following sports related concussion.

*What this study adds:* There is a subset of retired athletes who are better able to withstand the cognitive impact of sports related concussion given higher cognitive reserve.

*How this study might affect research, practice or policy:* Future research needs to explore modifiable factors to protect brain health in retired athletes, with the use of clinically meaningful measures of cognitive functioning. There is a need for longitudinal assessment and follow-up to clarify the role of cognitive reserve in retired athletes.

## INTRODUCTION

Traumatic brain injury, which includes concussion, is a risk factor for dementia (1) and early death (2). Cognitive reserve has been hypothesised as a protective mechanism against the expression of the cognitive effects of brain injury. Individual differences in cognitive performance or neural networks underlying task performance may allow some to cope better with the effects of brain injury and reduce the risk of dementia (3). Measures of cognitive reserve include estimated premorbid levels of intellectual function and education. Cognitive reserve is not a fixed process and derives from factors including education, occupation and leisure pursuits which suggests possible steps to reduce dementia risk.

There is growing interest in the long-term effects of concussion and/or repetitive head trauma, notably in athletes with a career in contact sports. The results of retrospective studies into long-term impacts of sport-related concussion on cognitive function are mixed. Cunningham et al. (4) reported declines in memory, executive functioning and processing speed; Hart et al.(5) reported cognitive deficits in naming and memory in retired athletes; and Pearce et al. (6) reported impairments in reaction time, paired associate learning, spatial working memory and cognitive flexibility in retired rugby league players relative to age and education matched controls. Guskiewicz et al. (7) reported that the incidence of reported symptoms of Mild Cognitive Impairment (MCI) was higher in retired football players relative to the general population.

However, other studies have not found clinically meaningful changes in cognitive performance. McMillan et al. (8) reported lower scores in a concussed group of retired athletes relative to controls, however these remained in the normal range. No relationship was found between frequency of concussion and cognitive function. In a comparison of retired hockey players and age matched controls, concussion history was inversely related to performance on intellectual/executive function measures (9) but the authors could not rule out the contribution of mood and other non-concussion factors. Similarly, Willer et al. (10) reported no difference in neuropsychological performance between retired NFL and National Hockey League players and controls.

Equestrian sports have a high level of concussive injury (11). The current study investigated the long-term neuropsychological outcome in a sample of retired, predominantly equestrian athletes compared to age and gender-matched controls using neuropsychological measures, and whether pre-morbid cognitive reserve (3) has a protective effect against cognitive impairment. This study is part of a larger study long-term study of neurodegenerative disease in the setting of brain trauma - the International Concussion and Head Injury Research Foundation (ICHIRF)-BRAIN study.

## METHODOLOGY

### Participants

Recruitment was via publicity in sports related media asking for volunteers with a history of concussion and control subjects to enrol. 1200 Interested volunteers completed an online questionnaire regarding self-reported concussion history, mood, sleep and physical and mental health status. 787 completed questionnaires were included in the final database. 318 volunteers were invited to attend a screening day, of which 32 declined. 166 were finally screened before Covid-19 prevented travel.

Number of concussions was checked during the initial interview. The nature of the competitive sport was varied and included equestrian, horse racing, motorsports, rugby, soccer and boxing.

Inclusion criteria: Participants were eligible to participate if they (i) were retired from their primary sport; (ii) were able to complete the online screening assessment; (iii) could understand and participate in the testing procedures; and (iv) were able to provide informed consent. Exclusion criteria were: being aged < 18 years; a history of self-reported previous severe traumatic brain injury; currently taking sedative or psychotropic medication; or a pre-existing medically diagnosed neurological disorder (e.g., dementia or Parkinson’s).

The aim of the study was to examine the long-term neuropsychological function of retired athletes with a history of sport-related concussion. We hypothesised that retired athletes with a history of concussion would have impaired neuropsychological functioning relative to matched controls, and that cognitive reserve would have a protective effect against deterioration.

## PROCEDURE

Ethical approval was obtained from St Mary’s University, London, on 1st June 2015, SMEC_2015-16_53, SMEC_2016-17_115 and SMEC_2017-18_051. All participants provided written informed consent. The trial was registered as ISRCTN 11312093.

### Outcome Measures

The neuropsychological tests selected aimed to capture change in a broad range of cognitive domains including language, memory, speed of processing and executive functioning. The test battery was developed with consideration of tests used in the long-term follow-up of athletes (12); tests previously used in the assessment of concussion in active jockeys (11); and of the likely older age of participants.

The domains assessed were: premorbid function (Test of Premorbid Function; TOPF (13)); vocabulary and verbal ability (Vocabulary subtest from Wechsler Adult Intelligence Scale-Fourth Edition; WAIS-4 (14)); auditory verbal short term and working memory (Digit Span subtest from the WAIS-4); processing speed (Symbol Digit Modalities Test; SDMT (15); and Speed of Comprehension Test (16)); verbal learning and memory (California Verbal Learning Test II; CVLT2 (17)); response inhibition (Stroop (18)), visual scanning and response alternation (Colour Trails Test (19)); and verbal fluency using category and letter paradigms.

All neuropsychological tests were administered and scored according to standardised instructions by the same qualified Clinical Neuropsychologist, blinded to the participant’s grouping of concussed or control.

### Statistical Analysis

A Principal Components Analysis (PCA) was conducted on 14 test scores as follows: TOPF; Vocabulary raw score; Digit span total; Symbol digit total; CVLT2 total, short delay free recall total, short delay cued recall total, long delay free recall total, long delay cued recall total and Recognition hits; FAS total; Animals total score; Stroop colour word number correct; Trail time 1 and 2 (in seconds). Outliers were explored using boxplots (Supplementary Figure S1) and a linear relationship between all variables was observed, though the limited range of some scales (e.g. Recognition hits, Stroop colour word number correct) led to a bulk of data points clustered at the limits. Scale consistency was achieved using normalisation, centring all variables to be distributed around a mean of 0 with standard deviation of 1. Principal components that in combination explained >95% of the variability in the included variables were selected for further exploration and analysis using linear regression.

To explore the association between test score components (dependent variables) and premorbid test scores (independent variable) in both concussed and control participants, principal component linear regression was used. This allowed for exploration of potentially confounding patient characteristics. Additional independent variables for age, gender and handedness (right- or left-handed) were explored in the model to adjust for potential confounding. To identify an appropriate model with the best statistical fit, covariates were added in a forward stepwise selection, using likelihood ratio tests to compare between nested models. Non-linear (quadratic) terms were explored for continuous variables, and interaction terms were tested between all variables included in the model.

Within the analysis, violations of normality were explored using normal quantile Q-Q plots. Where normality was found to be violated, different transformations of the PCA components were explored to resolve this. Homoskedasticity of residuals was tested using a Breusch-Pagan test (20). Where this was found to be violated, model standard errors were estimated using robust sandwich variance estimation (21). Potentially influential outliers were identified by plotting the residuals from final models.

## RESULTS

### Demographics

**Table 1:** Patient characteristics in the dataset.

|  | **Total (N=149)** | **Concussed (N=118)** | **Controls (N=31)** |
| --- | --- | --- | --- |
| Male (%) | 53 | 53.4 | 51.6 |
| Age (mean, range) | 61.4 (43.8, 79.8) | 61.5 (43.8, 79.8) | 61.1 (49.7, 74.3) |
| Years of education (median, range) | 13 (9, 19) | 13.5 (9, 19) | 13 (10, 19) |
| Premorbid functioning (median, range) | 61 (22, 69) | 60 (23, 69) | 63 (22, 69) |

166 participants were included in the dataset. Of these, 11 were missing data on at least one neuropsychological test and so were excluded from the dataset. Four participants were found to have high residual values n preliminary models fitted, and they had also previously been identified as having additional problems that affected their test scores. Two participants were also identified as having extreme/implausible test scores for CVLT2 and FAS. All six participants were excluded. This left 149 participants remaining for a complete-case analysis. There was no significant difference between the ages (t = 0.63, p = 0.53); years of education (t = -0.51, p = 0.0.6) and estimated premorbid level of functioning between the concussed and control groups (t = -1.33, p = 0.2).

The PCA indicated that three components cumulatively explained >95% of total variance, and 87% of variance in test scores. Principal component screeplots and biplots are shown in Figures S2 and S3 respectively. All variables apart from digit span total and FAS total have high loadings for PC1, though loadings for Trail 1 and 2 times are negative, whilst for other variables it is positive. This supports the negative correlation between these sets of variables (Figure S4). For PC2, loadings are strongly positive for digit span total and FAS total, and are negative for CVLT2, recognition hits, and all short and long delay free and cued recall. Other variables have less strong loadings for PC2. FAS and digit span total are the only variables with significant contributions to PC3, with positive and negative loadings respectively.

Preliminary regression checks showed that premorbid functioning was similar between concussed and control participants, and that premorbid functioning remained stable over age within the dataset.

The selected PC1 regression model had a multiple R2 of 0.541 (so explaining 54% of variance in PC1 values). The Breusch-Pagan test indicated significant heteroskedasticity (p=0.020), and so standard errors for model coefficients were adjusted using robust sandwich variance estimation. The interaction between premorbid function and concussion was statistically significant for a Bonferroni-adjusted alpha value of 0.017 (p<0.001). Model coefficients are shown in Table 2. The association between premorbid function and PC1 for a male aged 61 (the median age in the dataset) is shown in Figure 1.

**Table 2:**
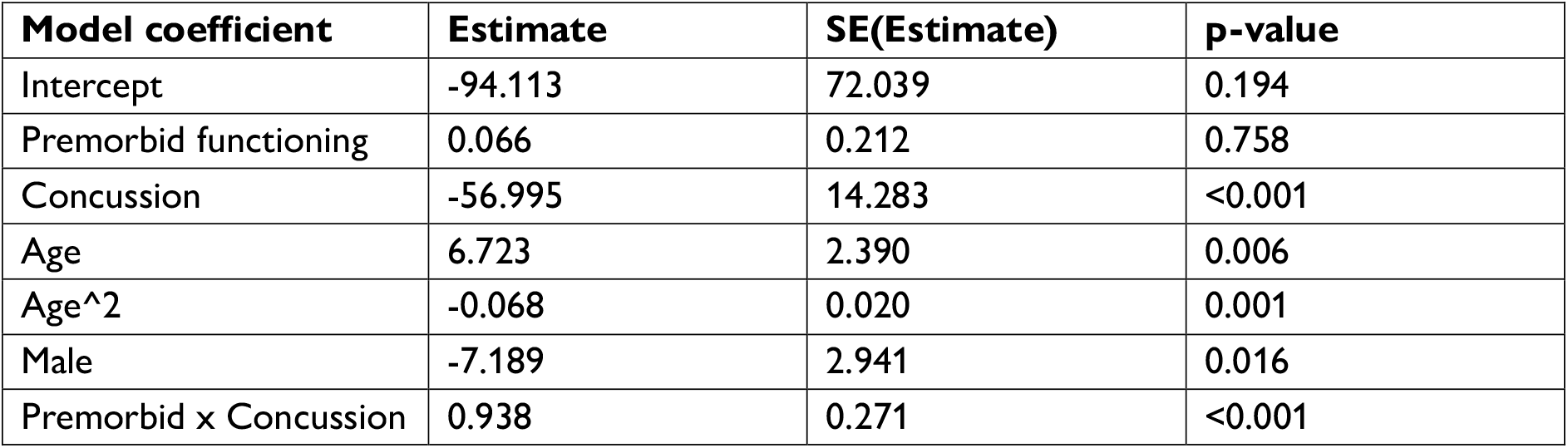
Model coefficient estimates and p-values for the selected model for PC1. Adjusted R2 = 0.522.

**Figure 1:**
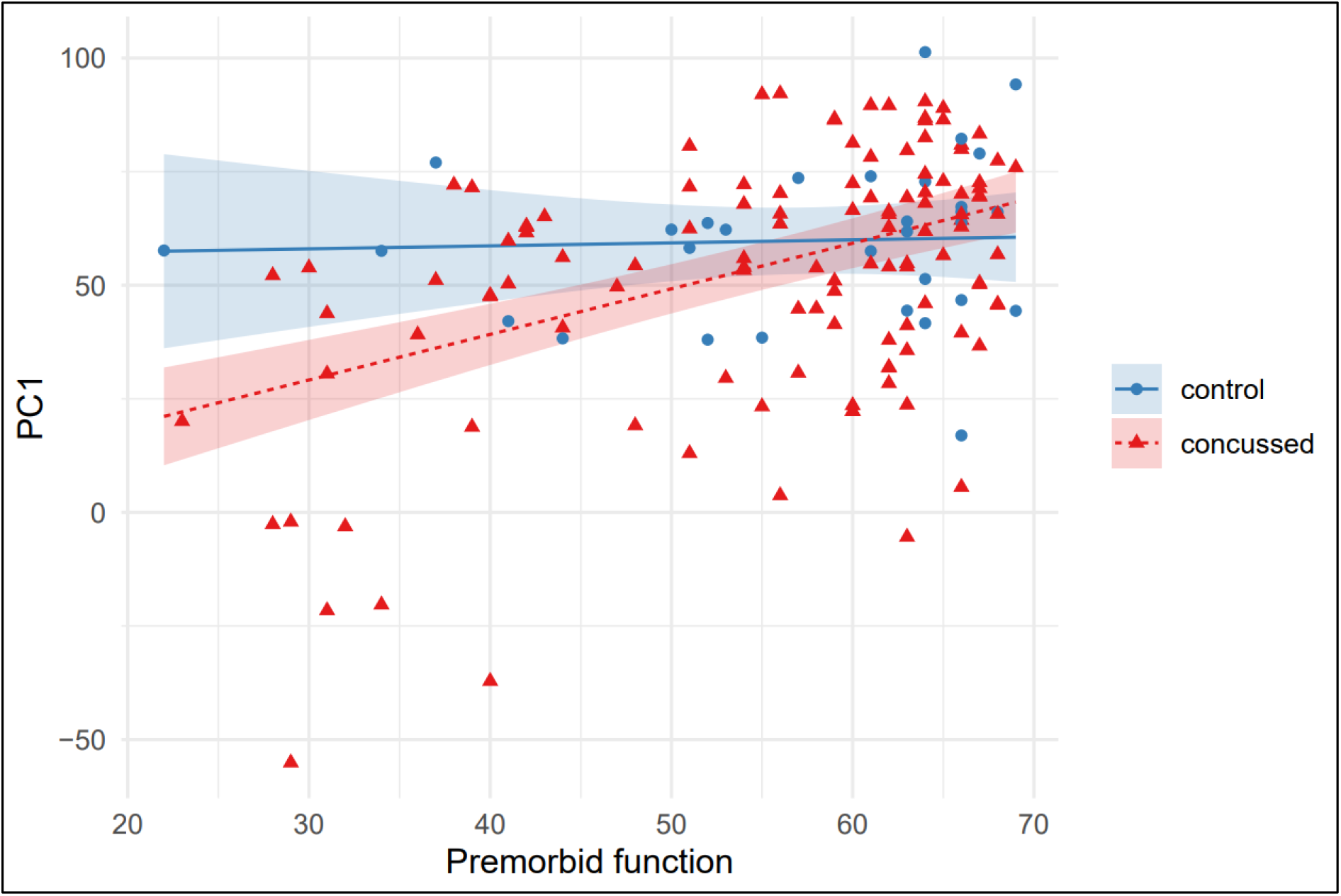
PC1 predictions from the selected regression model plotted against premorbid function for a male aged 61 (lines) and 95%CI (shaded regions), overlaid on the raw data for concussed (red) and controls (blue). 95%CI are calculated assuming homoskedasticity of residuals and so may be slightly wider than model coefficients, for which robust sandwich variance estimation is used.

The different gradient of the linear predictor for concussed and control participants indicates the impact of the interaction and implies that whilst there is a strong association between lower premorbid function and lower PC1 in concussed participants, control participants have similar PC1 regardless of premorbid function.

For PC2 the selected model included covariates for premorbid function, concussion, age, a quadratic (non-linear) term for age, and the interaction between premorbid function and concussion. The model had a multiple R2 of 0.448. The different gradient of the linear predictor for concussed and control participants indicates the impact of the interaction and implies that whilst there is a strong association between lower premorbid function and lower PC2 in concussed participants, this association is much less in control participants (as shown in Figure S5). However, unlike for PC1, there is some trend (non-significant) towards lower PC2 in control participants with lower premorbid function.

For PC3, the inclusion of concussion exposure as a covariate was not significant (p=0.379), and neither was the inclusion of exposure with the interaction between exposure and premorbid function (p=0.595) as shown in Figure S6.

## DISCUSSION

The interaction between concussion and premorbid functioning for the first and second components of the PCA indicates a strong association between lower premorbid functioning and lower neuropsychological functioning in concussed participants that is not present in control participants. Overall, this lends support to the hypothesis that cognitive decline due to concussion is dependent on the amount of cognitive reserve, with those with a higher cognitive reserve experiencing less cognitive decline. Premorbid functioning scores were found to be similar between concussed and control volunteers, and also remained stable over age within the dataset. That is, concussion did not have an effect on the TOPF which validated the use of this score as a stable measure of cognitive capacity and supports the other findings of the analysis.

PC1 explained 86.5% of the variance of the variables included in the PCA, and as is the most important component for interpreting results and considering the impact of concussion on cognitive decline. This component explained test scores well, indicating that it is a useful approach to reduce dimensionality in this dataset. The observed interaction between concussion and premorbid test function for the component implies that concussed participants with lower premorbid functioning have lower neuropsychology test scores in all domains.

For PC2, the loadings of individual neuropsychology tests are more complex, and the proportion of variance explained (6.7%) is smaller, making interpretation more challenging and less relevant for exploring cognitive decline. A smaller interaction was observed for PC2 than for PC1, but the findings broadly replicate those seen for PC1. For PC3 no significant interaction was observed and as this only explains 2.6 % of the variance it suggests minimal clinical significance.

### Cognitive reserve as a protective factor against cognitive deterioration

In the current study, a history of concussion was associated with lower neuropsychological test scores and this effect was more pronounced in those with lower cognitive reserve. The implications of low neuropsychology scores within this population is not clear. It may reflect a static condition or may reflect the possibility of further cognitive decline in keeping with MCI. Guskiewicz et al. (7) reported a higher incidence of MCI in a sample of NFL players. Increased mortality from neurodegenerative disease was found in a sample of retired professional soccer players and dementia-related medications were prescribed more to former players than controls (22).

Our findings suggest that cognitive reserve protects against cognitive decline associated with a history of sport-related concussion. Longitudinal assessment will determine whether cognitive reserve protects completely against deterioration or only delays the onset of cognitive changes relative to controls with no history of brain trauma. Broglio et al. (23) have suggested that there is a subset within the population able to sustain a concussion without clinically meaningful decline because of their reliance on cognitive reserve, consistent with our findings. Conversely, it has been reported that brain injury acts to accelerate cognitive decline and bring forward the age of dementia onset (24). Assessment of retired athletes at a more than one time point will determine if there is progressive deterioration and the rate of any decline.

Cognitive reserve is not static. Greater reserve may allow an individual to compensate for cognitive changes (3) or access resources to maintain everyday functioning. Potentially modifiable risk factors for dementia include education, drug and alcohol consumption, level of daily activity, social isolation, and comorbid medical conditions (25-27). It may therefore be possible to adopt a preventative approach in active athletes and identify those at greatest risk and encourage further education, a healthy lifestyle and resilience to boost cognitive reserve and shield against the cognitive decline associated with concussion. Walton et al. (28) identified that the combined beneficial effects of exercise, sleep and better diet was similar or greater than the negative effects of concussion history on self-reported cognitive and mood changes in a sample of retired NFL players. Exploration of modifiable factors was beyond the scope of the present study. Longitudinal follow-up is required to better define possible trajectories following sport-related concussion and determine the expression of cognitive impairment in individuals with lower cognitive reserve as risk factors are modified.

### Cognitive outcome following sport-related concussion

Other retrospective studies in retired athletes have produced mixed results regarding evidence of cognitive impairment following sport-related concussion. De Beaumont et al. (29) found reductions in episodic memory and response inhibition in retired athletes with a history of sports concussion and suggested that this was due to a lower inability to compensate for demands on attentional resources. In a comparison of retired hockey players and controls, concussion history was inversely related to performance on intellectual/executive function (9), although the contribution of education and substance abuse could not be dismissed. In contrast, McMillan et al. (8) did not find a relationship between the frequency of concussion and cognition in a sample of 52 retired rugby players. In a study of 30 retired Australian Football players who were re-administered a battery of neuropsychological tests 25 years after initial assessment, concussion history was not found to influence cognitive or psychological functioning and there was no change in cognitive function over time (12). Willer et al. (10) compared the neuropsychological performance of retired football and hockey players with both non-contact sport athletes and non-sport controls to find no difference between their performance and that of the control groups. The negative findings in these studies may reflect smaller sample sizes or the contribution of non-concussion-related variables.

### The contribution of non-brain injury factors

The results of the current study indicate an association and further work is required to support a causal relationship. Characteristics related to retired athletes may have had an impact on the findings and future studies need to take these factors into account. The contribution of non-trauma-related variables to neuropsychological test performance are highlighted in studies of retired athletes. Casson et al. (30) examined retired NFL players and found that 24% of the sample had one to two impairments on cognitive tests. Non-trauma-related variables, including alcohol intake and body mass index were associated with as many if not more abnormal scores than sport-related factors. Comorbidities including mood disorder, steroid use (31), history of learning difficulties (32), drug and alcohol history, pain and other physical comorbidities (8) are suggested as contributing factors to subjective reports of everyday difficulties and/or impairment on formal cognitive assessment in retired athletes. It is a limitation of our study that non-trauma factors were not controlled. Future longitudinal studies are required to determine the influence of these non-concussion factors on neuropsychological outcomes.

### Strengths and limitations

The strength of the current study is the relatively large sample size to specifically examine the neuropsychological functioning of retired athletes with the administration of a range of tests.

However, it is important to note that the relationship observed here is correlational, and further work would be needed to support a causal relationship. Although the analysis adjusts for some key patient characteristics that may be confounders, there are potentially many others that could be important. Characteristics associated with cases versus controls may have a significant impact on the findings. For example, there may be numerous confounders (e.g. smoking, drinking) associated with profession (jockeys versus non-jockeys) that cannot be accounted for in this dataset.

Being able to characterise the observed association as a “decline” is challenging given that data at only a single point in time is available for each participant. Our findings therefore strongly depend on the validity of TOPF as being a stable measure over time (irrespective of cognitive decline). Additional follow-up measurements on the same participants would allow further exploration/confirmation of a cognitive decline within this dataset.

## Conclusion

In summary, the PCA has provided a robust summary measure of neuropsychological performance (PC1) which can be utilised in future studies of retired athletes, providing a practical brief test battery in an ageing cohort. Our results indicate that lower test scores are associated with sport-related concussion. The findings suggest a protective effect of cognitive reserve on long-term cognitive changes associated with a history of sport-related concussion in a population of retired athletes functioning normally in society.

## Data Availability

All data will be available at the end of the 10 years study 2015-2025 upon reasonable request to the authors

## ACKNOWLEDGEMENTS

The ICHIRF-BRAIN Study would not be possible without the input of the ICHIRF Project Manager, Pippa Theo.

## DISCLOSURE

Michael Turner is employed as CEO and Medical Director of the International Concussion and Head Injury Research Foundation (ICHIRF) and was formerly employed as the Chief Medical Adviser to the British Horseracing Authority (BHA) and the Lawn Tennis Association (LTA). He is Honorary Medical Adviser to the Professional Jockeys Insurance Scheme (PRIS) for which he receives a discretionary honorarium. ICHIRF is a not-for-profit organisation, and the donors are listed elsewhere in the document. He undertakes no clinical duties but has been reimbursed for travel and accommodation at conferences, symposia and scientific meetings by the organisers. He does not hold any shares in any company related to concussion or brain injury assessment or technology.

## FUNDING INFORMATION

The ICHIRF project is currently philanthropically funded by Godolphin Racing, the Injured Jockeys Fund (UK), the Irish Injured Jockeys Fund, the Players Foundation, the National Football League (US), the Concussion Foundation, the Racing Foundation, the British Association of Sport and Exercise Medicine as well as private donations.

## DATA STATEMENT

No data available

## PATIENT AND PUBLIC INVOLVEMENT

There was no patient involvement in the development of the research.

## AUTHOR CONTRIBUTION STATEMENT

Bonnie-Kate Dewar: Conceptualisation, Methodology, Validation, Resources, Investigation, Writing-Original draft preparation, Writing-Review and editing

Hugo Pedder: Formal analysis, Investigation, Methodology, Software, Visualization, Writing - Original draft preparation, Writing - Review and editing.

James Murray Conceptualization; Methodology; Writing - Review & Editing.

Michael Turner Conceptualisation, Resources, Review and editing, Project administration, Funding

## Notes

### Competing Interest Statement

The authors have declared no competing interest.

### Author Declarations

Ethical approval was obtained from St Marys University Twickenham Waldgrave Road London TW1 4SX UK on 1st June 2015 SMEC_2015-16_53 SMEC_2016-17_115 and SMEC_2017-18_051. Ethical approval was obtained from St Marys University, United Kingdom. All participants provided written informed consent. The trial was registered as ISRCTN 11312093. SMEC= St Marys' Ethical Committee

