## Supplementary material for "LONG TERM NEUROPSYCHOLOGICAL OUTCOME AND EFFECT OF COGNITIVE RESERVE IN RETIRED ATHLETES": Figures S1 to S6

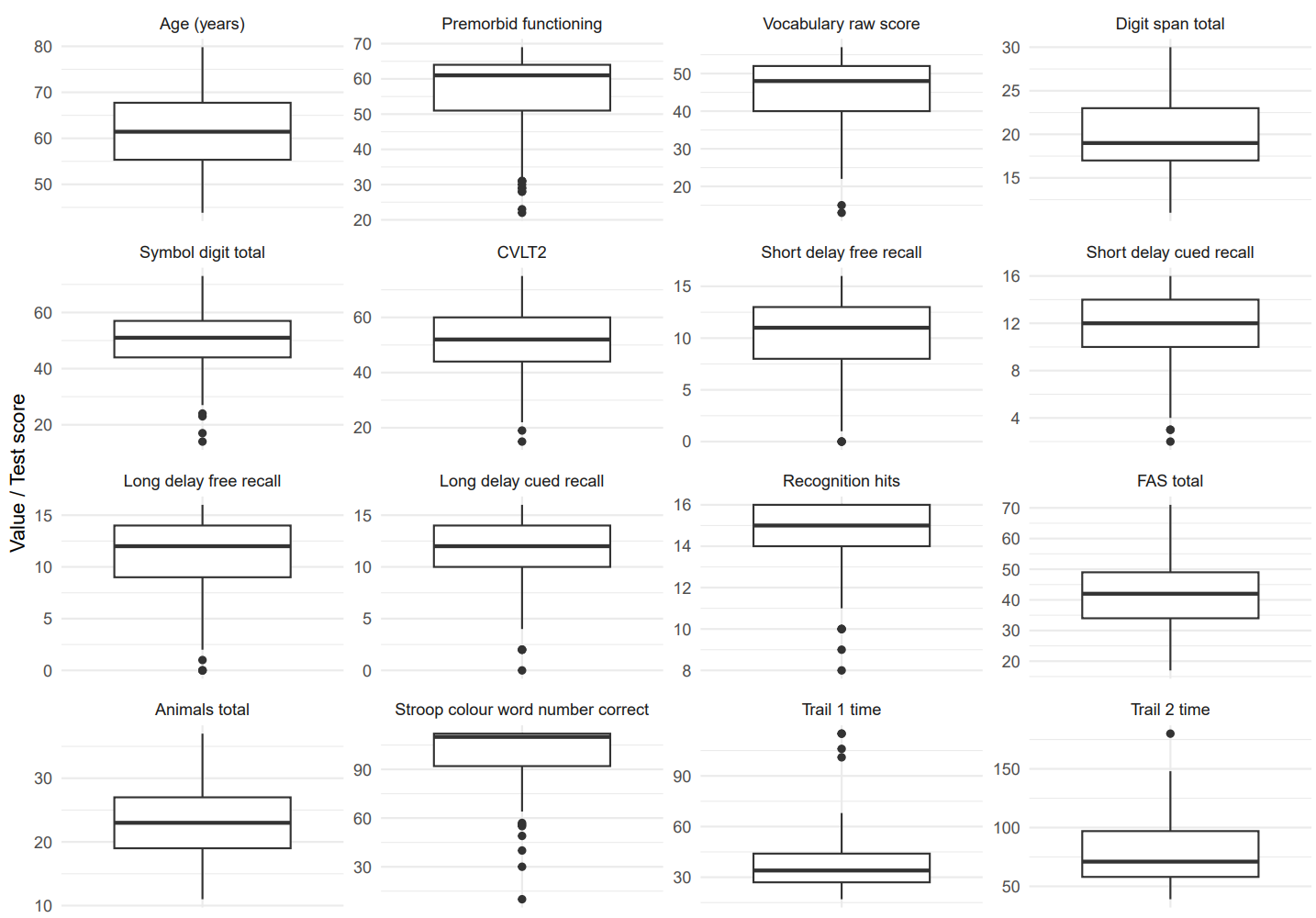

Figure S1: Boxplots of continuous variables included in the dataset.

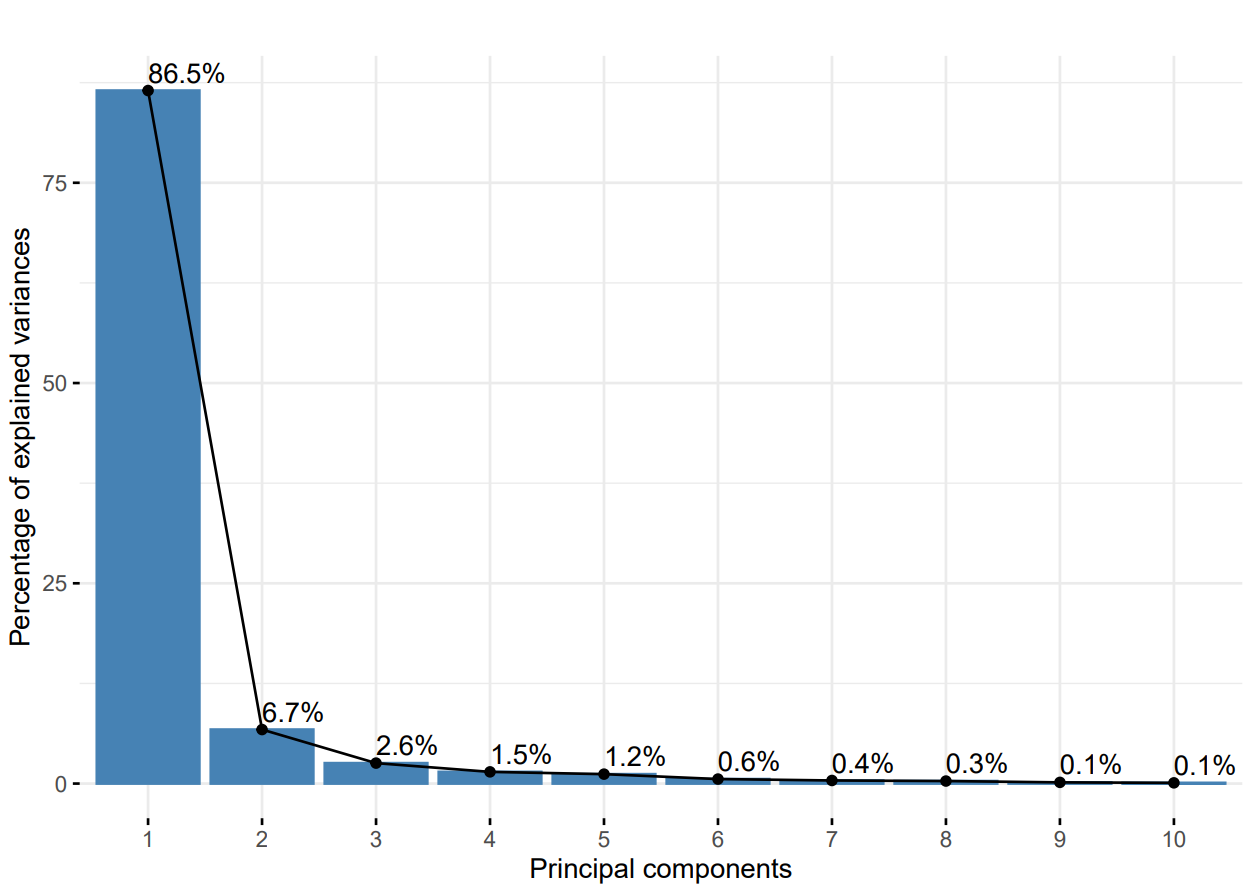

Figure S2: Principal Component Analysis (PCA) screeplot showing the proportion of the total variance explained by each Principal Component across 14 variables included in the analysis.

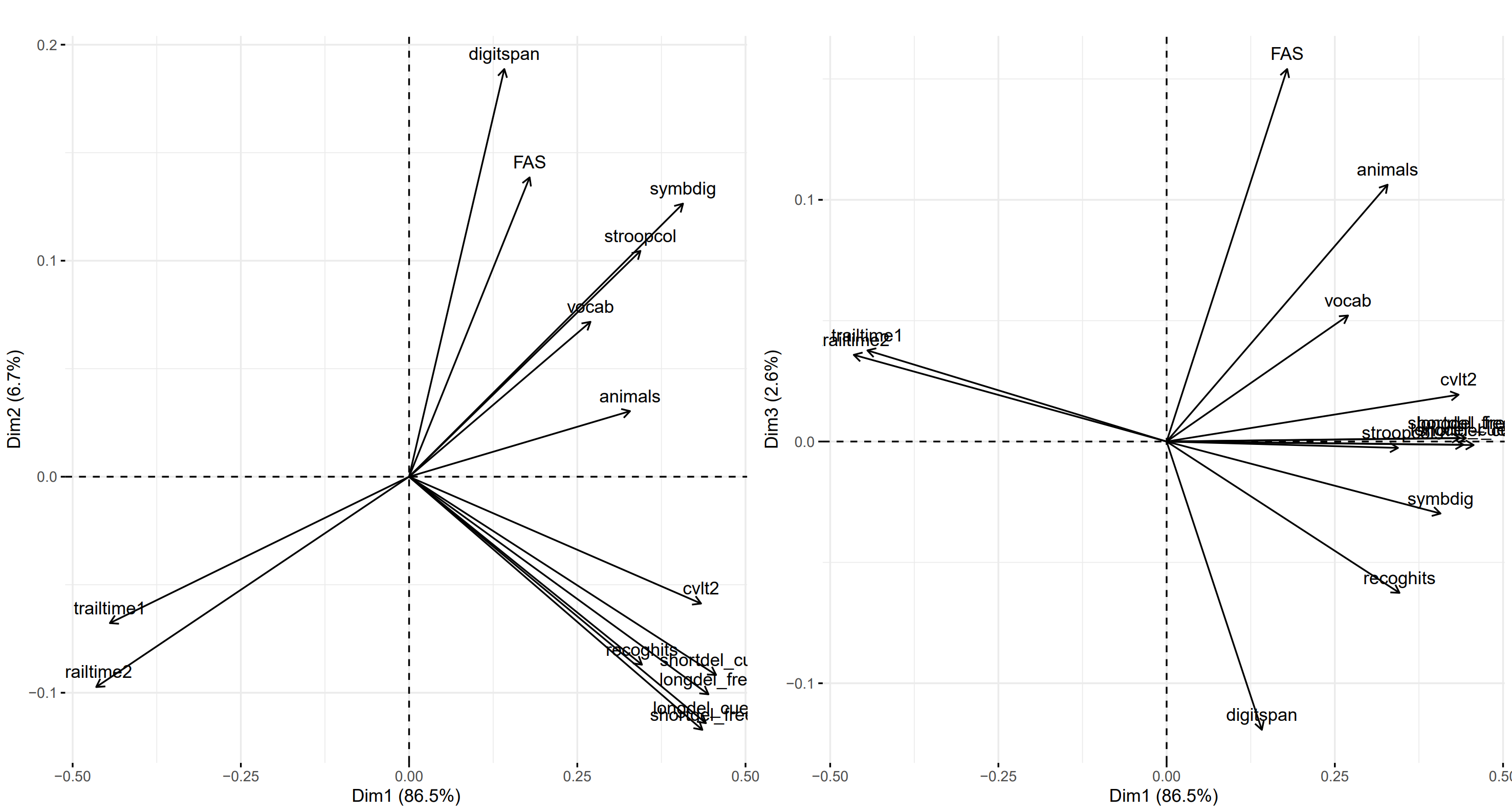

Figure S3: Principal Component Analysis (PCA) biplots. These indicate the loadings for variables on PC1 and PC2 (left panel) and on PC1 and PC2 (right panel). Variables grouped together more closely are more positively correlated, whilst those at opposite directions from the origin are more negatively correlated. Greater distances from the origin represent higher loadings, which indicates better representation within a particular Principal Component.

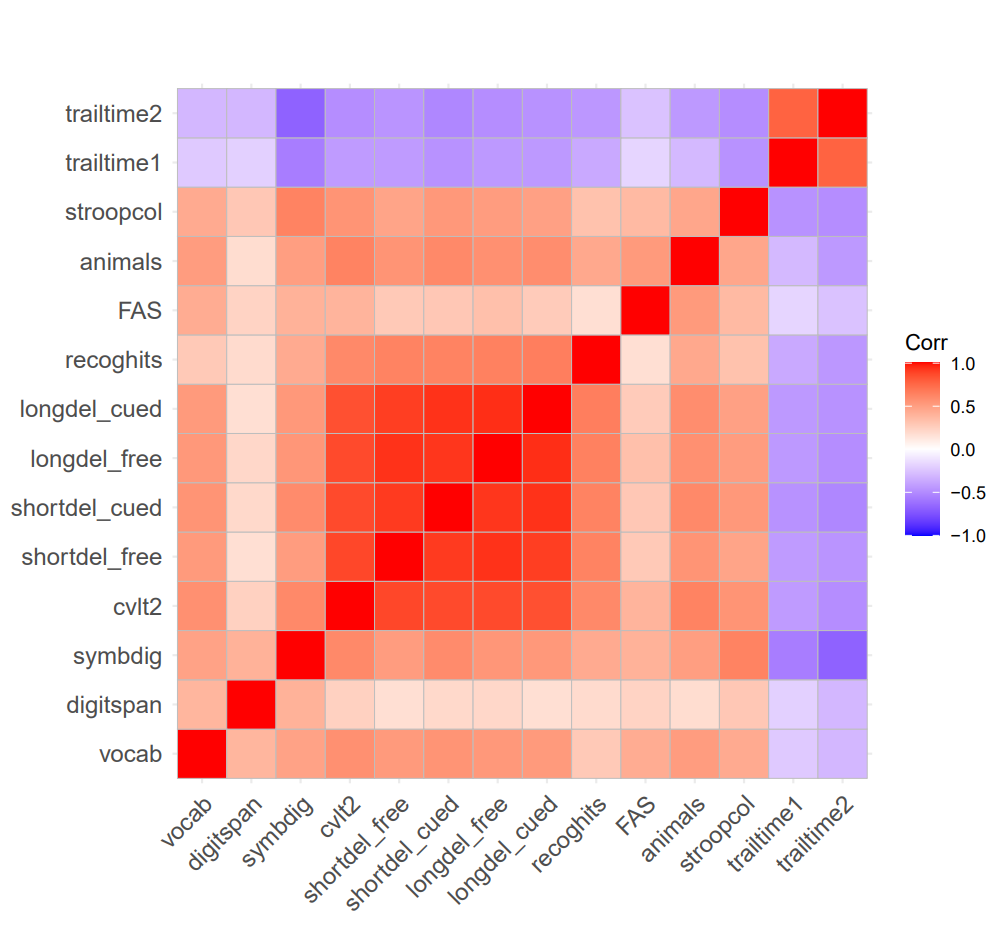

Figure S4: Correlation matrix between variables included in PCA. Red shading indicates variables that are positively correlated and blue shading indicates those that are negatively correlated, with darker shading indicating stronger correlation. trailtime1 = Trail 1 time, trailtime2 = Trail 2 time, stroopcol = Stroop colour word number correct, animals = animals total score, FAS = FAS total score, recoghits = recognition hits, longdel_cued = long delay cued recall, longdel_free = long delay free recall, shortdel_cued = short delay cued recall, shortdel_free = short delay free recall, cvlt2 = CVLT2 total score, symbdig = symbol digit total score, digitspan = digit span total score, vocab = vocabulary raw score

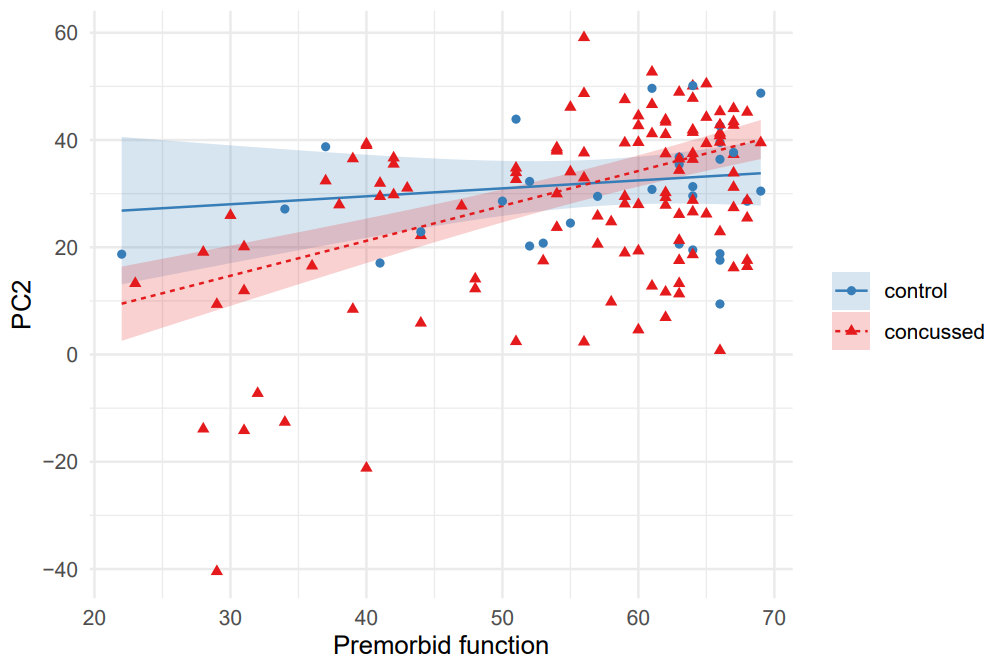

Figure S5: PC2 predictions from the selected regression model plotted against premorbid function for 61-year old (lines) and 95%CI (shaded regions), overlaid on the raw data for concussed (red) and controls (blue). 95%CI are calculated assuming homoskedasticity of residuals and so may be slightly wider than model coefficients, for which robust sandwich variance estimation is used.

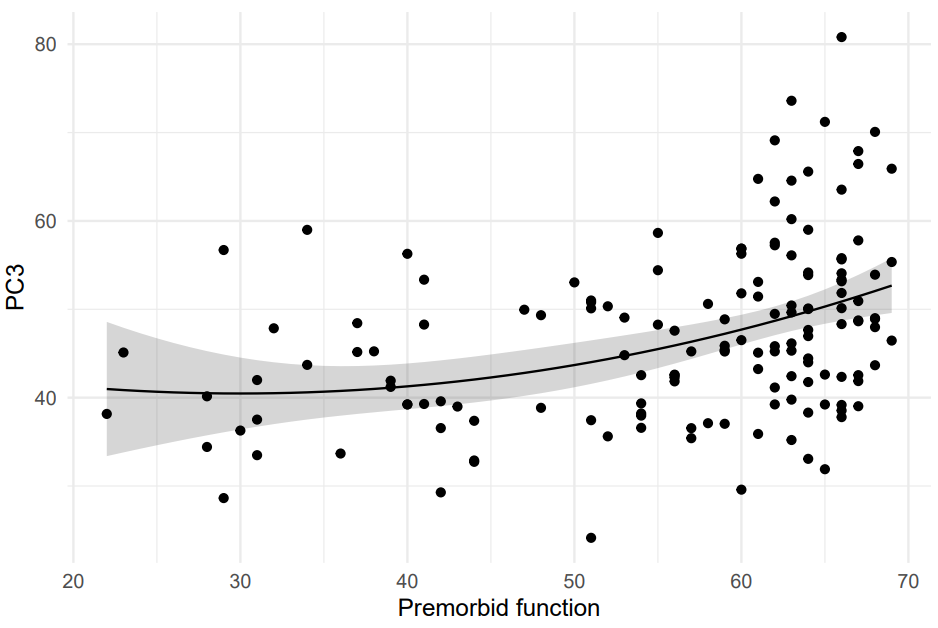

Figure S6: PC3 predictions from the selected regression model plotted against premorbid function for a 61-year old (lines) and 95%CI (shaded regions), overlaid on the raw data. 95%CI are calculated assuming homoskedasticity of residuals and so may be slightly wider than model coefficients, for which robust sandwich variance estimation is used.
